# Coronavirus disease-19: The First 7,755 Cases in the Republic of Korea

**DOI:** 10.1101/2020.03.15.20036368

**Authors:** COVID-19 National Emergency Response Center, Epidemiology Center, Epidemiology & and Case management team, Korea Centers for Disease Control and Prevention

## Abstract

We report the first 7,755 patients with confirmed COVID-19 in Korea as of March 13, 2020. A total of 66 deaths were identified, resulting case fatality proportion of 0.9%. Older people, and those with coexisting medical conditions were at risk for fatal outcomes. The highest number of cases were from Daegu, followed by Gyeongbuk, with elevated age-stratified case fatality. This summary may help to understand the disease dynamics in the early phase of COVID-19 outbreak, therefore, to guide future public health measures.

## Introduction

Only a few weeks after the first case of coronavirus disease 2019 (COVID-19) was noted in Wuhan, China, the first imported case of COVID-19 was confirmed in the Republic of Korea on January 20, 2020 (1). International travel has facilitated the spread of COVID-19 throughout the world. As of March 13, 2020, 118 countries, territories, and areas have reported COVID-19 cases, however, differences in patterns and intensity of transmission ranging were observed (2). In February and early March in Korea, a sharp increase in the number of COVID-19 cases was observed, with most infections in specific cluster and geographic region.

Based on the surveillance data, we report the basic epidemiological characteristics of COVID-19 in order to understand the early course of the pandemic. Furthermore, we investigated factors associated with deaths to provide evidence on vulnerable population for fatal outcome to guide public health prioritization.

## Materials and Methods

This is a summary of the first 7,755 patients with confirmed COVID-19 in Korea as of March 13, 2020. To obtain demographic, epidemiological, and early clinical information, COVID-19 reporting and surveillance data were retrieved from Korea Centers for Disease Control and Prevention (KCDC)-operated National Notifiable Disease Surveillance System (NNDSS) (3). Patient age was provided on the date of diagnosis, and key indicators such as estimated duration of exposure, date of onset of symptoms, and route of transmission were identified by field epidemiological investigators (1). It must be noted that the data presented in this summary may be changed depending on the results of further epidemiological investigation.

## Results

As of March 12, 2020, a total of 7,755 laboratory-confirmed cases of COVID-19 and 66 deaths were identified in Korea, resulting case fatality proportion of 0.9% (Table 1). The female-to-male ratio was 62:38. 20-29 years group accounted for 28.9% of all cases, followed by 50-59 years, and 40-49 years group. The case fatality proportion was 0.1% among 30-39 years and 40-49 years group, then increased to 0.4% (50-59 years), 1.5% (60-69 years), 5.0% (70-79 years), and 8.5% (≥80 years).

**Table 1.** Demography and case fatality proportion (CFP) of the first 7,755 laboratory-confirmed coronavirus diseases-19 cases, as of March 12 2020, Republic of Korea

| Variables | Confirmed cases |  | Deaths | CFP |
| --- | --- | --- | --- | --- |
|  | N | % | N | % |
| Sex |  |  |  |  |
| Female | 4,808 | (62.0) | 25 | (0.5) |
| Male | 2,947 | (38.0) | 35 | (1.2) |
| Age group (years) |  |  |  |  |
| 0-9 | 75 | (1.0) | - | - |
| 10-19 | 405 | (5.2) | - | - |
| 20-29 | 2,238 | (28.9) | - | - |
| 30-39 | 804 | (10.4) | 1 | (0.1) |
| 40-49 | 1,082 | (14.0) | 1 | (0.1) |
| 50-59 | 1,472 | (19.0) | 6 | (0.4) |
| 60-69 | 960 | (12.4) | 14 | (1.5) |
| 70-79 | 483 | (6.2) | 24 | (5.0) |
| ≥80 | 236 | (3.0) | 20 | (8.5) |
| Total | 7,755 |  | 66 | (0.9) |

The epidemic curves presented in Figures 1 and 2 are generated using National Notifiable Disease Surveillance System (NNDSS). Epidemic curve by date of onset precedes epidemic curve by date of diagnosis by few days or a week (Figure 1). It must be noted that the date of symptom onset may change depending on the results of further epidemiological investigation, and delayed reporting of cases. Figure 2 depicts epidemic curve by different regions of Korea: the nationwide, Daegu, Gyeongbuk, and Others (outside Daegu and Gyeongbuk). From mid-February an increased number of cases was reported with a peak in the late February and early March. Figure 3 shows the age distribution and sex ratio among Daegu, Gyeongbuk, and Others regions. Note the increased proportion of cases in 20-29 years and in females among cases in Daegu and Gyeongbuk.

**Figure 1.**
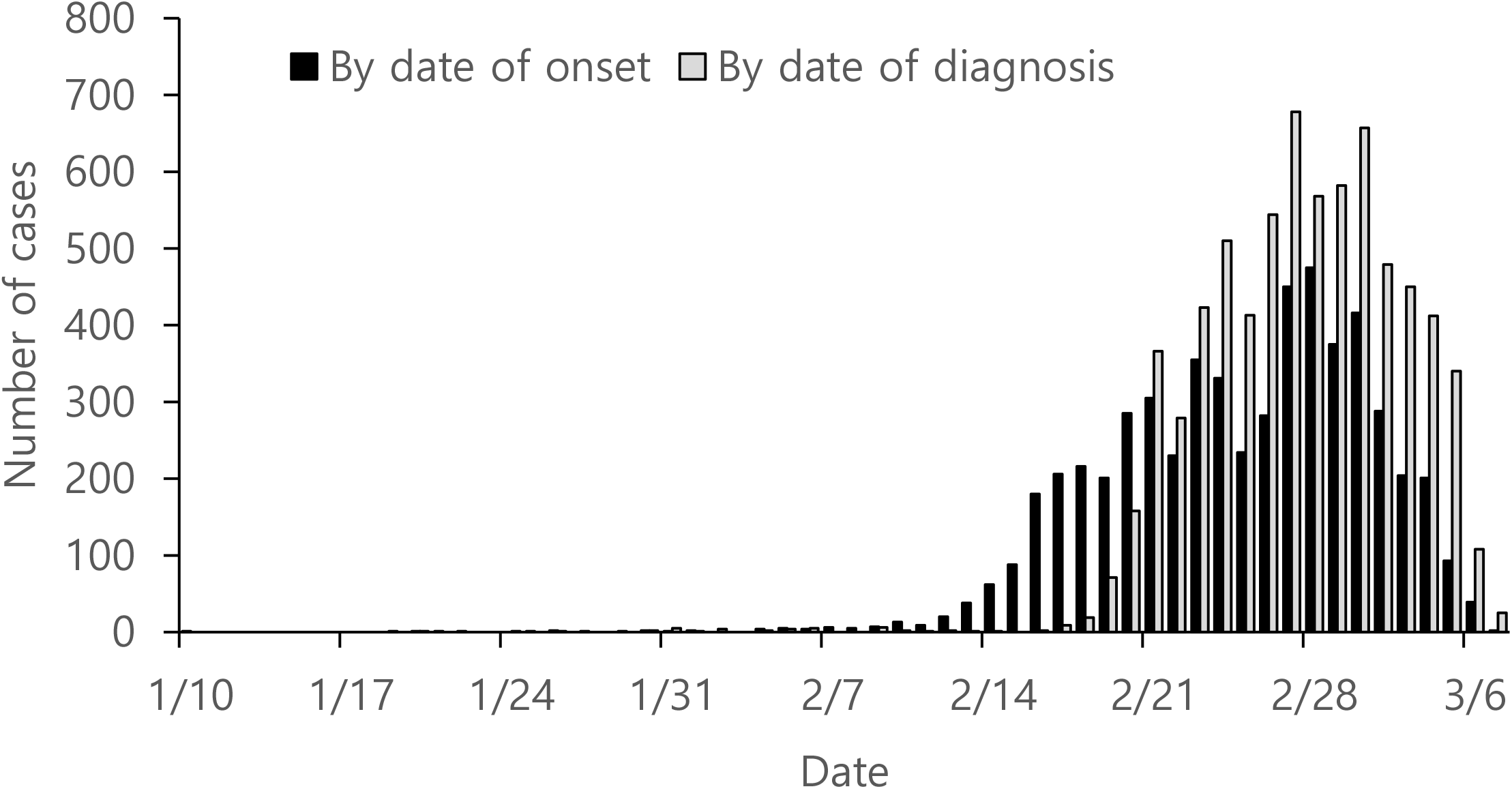
Epidemic curve of laboratory-confirmed coronavirus disease-19 by date of symptom onset and by date of diagnosis, as of March 7, 2020, Republic of Korea *Provisional data for date of symptom onset

**Figure 2.**
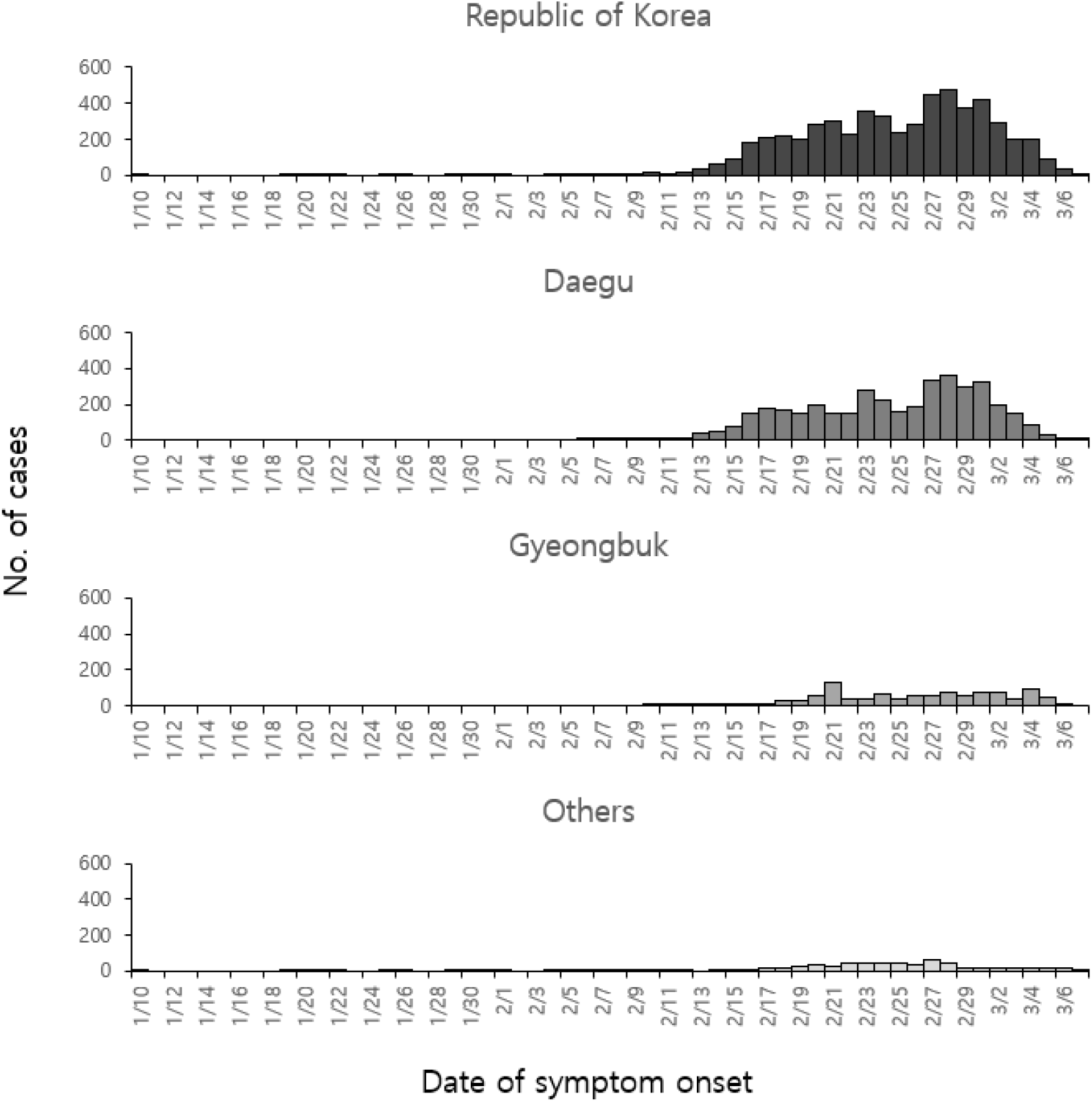
Epidemic curve of laboratory-confirmed coronavirus disease-19 cases by date of symptom onset, as of March 7, 2020, Republic of Korea *Provisional data for date of symptom onset

**Figure 3.**
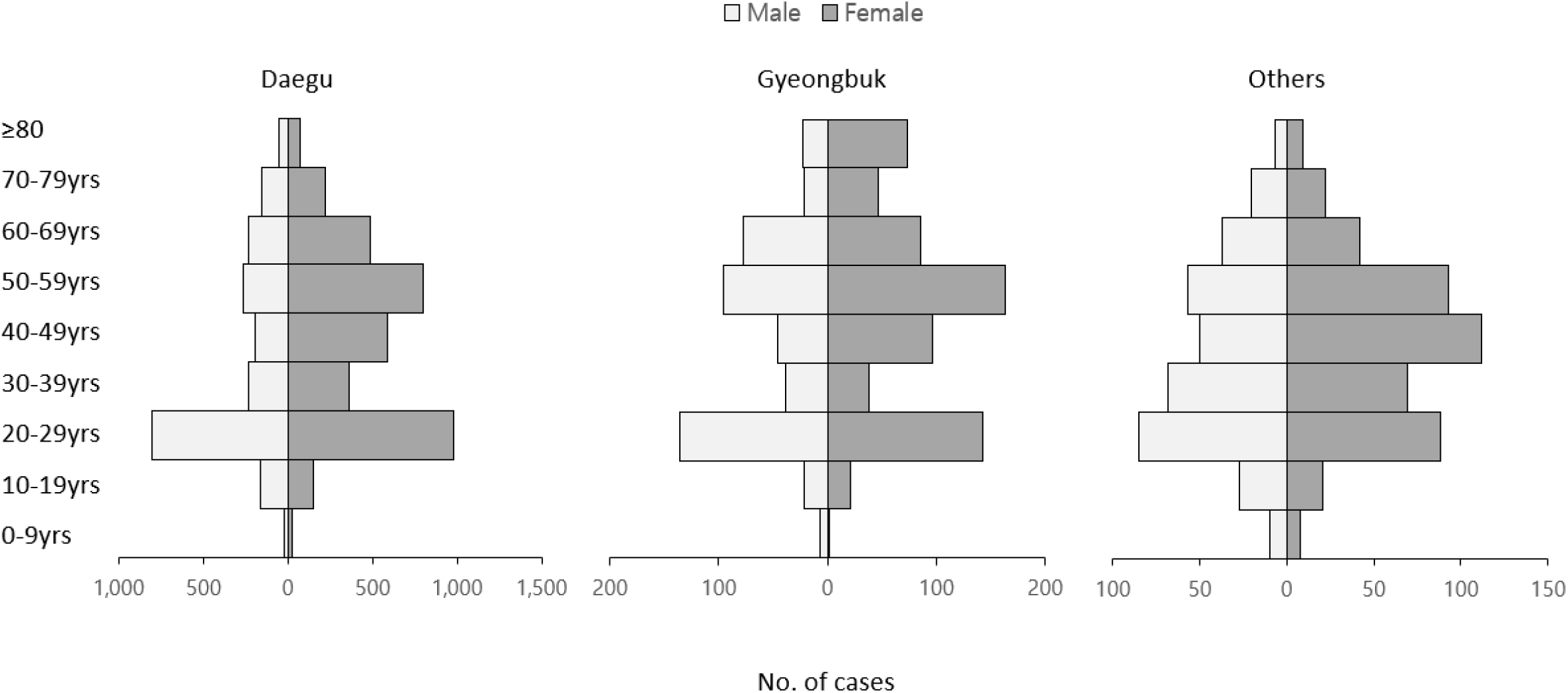
Age distribution and sex ratio of the first 7,755 cases of laboratory-confirmed coronavirus disease-19, as of March 12 2020, Republic of Korea

Case fatality proportion was the highest among persons aged ≥80 years in Daegu, followed by persons aged 70-79 in Daegu and elderlies in Gyeongbuk. Overall case fatality proportion in Daegu and Gyeongbuk were 0.8% and 1.4%, which were higher than that of Others regions with 0.4%. Figure 4 depicts age-specific epidemic curve stratified as 0-59 years and ≥60 years. The outbreak generally began with younger age group, followed by elderly population.

**Figure 4.**
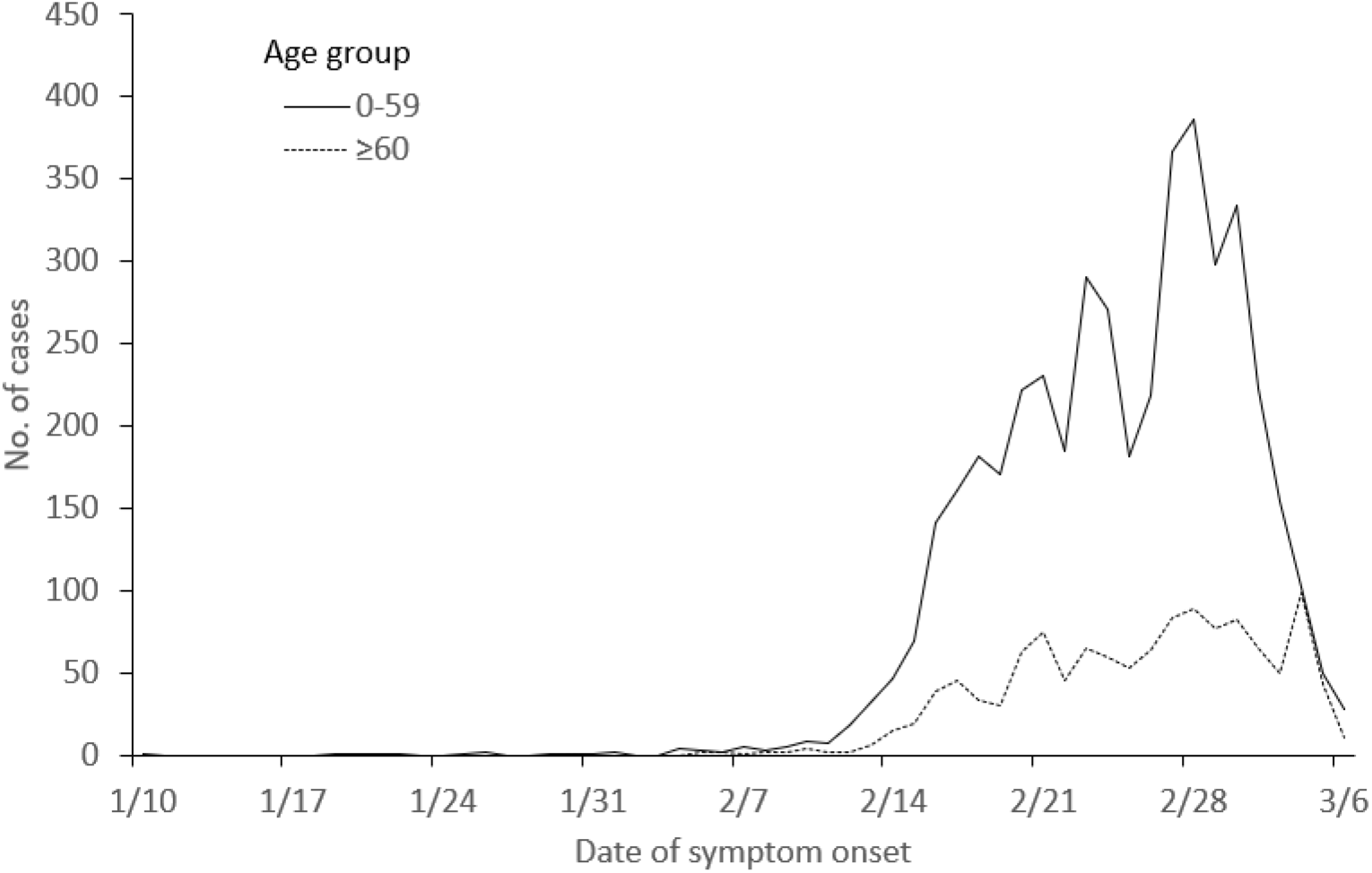
Age-specific epidemic curve of laboratory-confirmed coronavirus disease-19 cases by date of symptom onset, as of March 7, 2020, Republic of Korea *Provisional data for date of symptom onset

Table 3 describes the 66 fatal cases of laboratory-confirmed coronavirus diseases-19 cases, as of March 12, 2020. The median age is 77 years (range, 35-93 years), and female-to-male ratio of 44:56. Of 63 cases, 96.8% were found to have coexisting conditions: 47.6% hypertension, 36.5% diabetes, 16% neurodegenerative disorders, and 17.5% pulmonary diseases. 71.2% of fatal cases were from Daegu and 24.2% were from Gyeongbuk. 5 patients were succumbed to death at home, and 11 were diagnosed to have contracted COVID-19 after death. The median interval between onset of symptom and death was 10 days (range 1-24 days), while the median interval between date of hospitalization and date of death was 5 days (range 0-16 days).

**Table 2.** Number of deaths and case fatality proportion (CFP) of laboratory-confirmed coronavirus diseases-19 cases by geographic regions, as of March 12 2020, Republic of Korea

| Age group | Daegu |  |  | Gyeongbuk |  |  | Others |  |  |
| --- | --- | --- | --- | --- | --- | --- | --- | --- | --- |
|  | Deaths | Total cases | CFP | Deaths | Total cases | CFP | Deaths | Total cases | CFP |
| 0-9 | - | 48 | - | - | 9 | - | - | 18 | - |
| 10-19 | - | 314 | - | - | 43 | - | - | 48 | - |
| 20-29 | - | 1,787 | - | - | 279 | - | - | 173 | - |
| 30-39 | - | 590 | - | - | 76 | - | 1 | 137 | (0.7) |
| 40-49 | - | 778 | - | 1 | 142 | (0.7) | - | 162 | - |
| 50-59 | 1 | 1,061 | (0.1) | 4 | 260 | (1.5) | 1 | 150 | (0.7) |
| 60-69 | 10 | 719 | (1.4) | 4 | 162 | (2.5) | - | 79 | - |
| 70-79 | 21 | 373 | (5.6) | 3 | 69 | (4.3) | - | 42 | - |
| ≥80 | 15 | 124 | (12.1) | 4 | 96 | (4.2) | 1 | 16 | (6.3) |
| Total | 47 | 5,794 | (0.8) | 16 | 1,136 | (1.4) | 3 | 825 | (0.4) |

**Table 3.** 66 fatal cases of laboratory-confirmed coronavirus diseases-19 cases, as of March 12 2020, Republic of Korea

| Variables | N | (%) |
| --- | --- | --- |
| Age, years, median (range) | 77 | (35-93) |
| Sex |  |  |
| Female | 29 | (43.9) |
| Male | 37 | (56.1) |
| Coexisting condition* |  |  |
| Any | 61 | (96.8) |
| Cerebrovascular disease | 5 | (7.9) |
| Heart disease | 10 | (15.9) |
| Pulmonary disease | 11 | (17.5) |
| Kidney disease | 5 | (7.9) |
| Diabetes | 23 | (36.5) |
| Hypertension | 30 | (47.6) |
| Cancer | 7 | (11.1) |
| Neurodegenerative disorder | 16 | (25.4) |
| None | 3 | (4.8) |
| Under investigation | 3 | (4.5) |
| Region |  |  |
| Daegu | 47 | (71.2) |
| Gyeongbuk | 16 | (24.2) |
| Others | 3 | (4.5) |
| Death while in home | 5 | (7.6) |
| Diagnosis after death | 11 | (16.7) |
| Interval, days, median (range) |  |  |
| Symptom → Death | 10 | (1-24) |
| Symptom → Diagnosis | 4 | (0-20) |
| Symptom → Hospitalization | 4.5 | (0-11) |
| Hospitalization → Death | 5 | (0-16) |
| Total | 66 |  |

\* A single case may have multiple coexisting condition, proportion with only cases with investigation files

## Discussion

The analysis of the first 7,755 cases of COVID-19 in Korea showed a first increase of cases in specific geographic region comparable to the epidemiological situation in China and Italy (4, 5). In Korea, despite a relevant number of cases per week in late February and early March, the peak seemed to have levelled off, yet the first wave did not go away. With descriptive analysis of interim surveillance data, the data suggest that the patient outcome differed markedly between the geographic regions, highlighting the magnitude of surge capacity on access to hospitalitzation.

Summary of the epidemiological characteristics of the first 7,755 COVID-19 cases in Korea indicated that important differences could be observed between the geographic region affected by the magnitude of outbreak. Different factors might have played a role: triaging patients for hospitalization, social distancing reducing the number of contacts, and introduction of outbreak among vulnerable population. However, the strict containment strategy in the beginning of the COVID-19 importation phase may have contributed to lower wave in other parts of Korea. More severe cases were seen in elderly with coexisting condition, which warrants clear triage management and high-risk approach in healthcare access prioritization. The COVID-19 seemed not have resulted in fatal outcome among young adults and children, however their role in disease transmission should be studies in near future.

This summary may help to understand the disease dynamics in the early phase of COVID-19 outbreak, therefore, to guide future public health measures in other countries.

## Data Availability

Korea CDC

## Acknowledgments

We thank the relevant ministries, including the Ministry of Interior and Safety, Si/Do and Si/Gun/Gu, medical staffs in health centers, and medical facilities for their efforts in responding to COVID-19 outbreak.

